# Using Disability-Adjusted Life Years to assess the burden of road traffic injuries in Nay Pyi Taw, Myanmar

**DOI:** 10.1101/2020.04.26.20080325

**Authors:** Win Ei Ei Thaw, Wantanee Phanprasit, Kwanjai Amnatsatsue, Ariya Bunnagamchairat, Kanitta Bundhamcharoen

## Abstract

**Background:** Road traffic injury (RTI) is growing public health problem in Myanmar. In addition, every count in road crush is followed by remarkable burdens in the community. In this study, burden of disease (BOD) approach is used to quantify this hidden problem in the studied area.

**Aim/Objective/Purpose:** To assess the burden of traffic injuries in Nay Pyi Taw Union Territory, Myanmar

**Methods:** This cross-sectional study analyzed 16,338 traffic injury victims in injury registry, from 2012 to 2016. The burden was estimated by disability-adjusted life years (DALYs). The analysis was disaggregated by sex, age and type of road users, as well as expressed the rate by per 100,000 population.

**Results/Outcomes:** It was 60,962 DALYs in total for the studied period, with the rate of 1050.85 per 100,000 population. Although premature death in road crash was only 6.2% in the study, it attributed 87.8% of total burden. Total DALYs contributed by male were three-times higher than female. Nearly half of burden came from the young and productive population of 15-29 years. The highest DALYs rate was seen in the 20-24 years in male and 30-34 years in female. The highest RTI burden was due to motorized two-wheelers, with 69.4% of total DALYs.

**Conclusion:** This study initiates usefulness of local data from injury registry to calculate the burden of injury. The findings highlight a huge burden of traffic injuries in the community, focusing on the hidden contribution of fatal cases and the vulnerability of young adult, male and motorcyclists in traffic accidents.

## INTRODUCTION

In every country around the world, transport connected people to the opportunity; including education, health care and trade. Safe mobility was a basic right for all. Simultaneously, road traffic injury (RTI) was the major leading cause of death among the productive age group of 15 – 29 years in all regions,[1]. Ninety percent of traffic fatality occurred in developing countries,[1], assuming as a disease of development. The changing globalization, altogether with economic progress, urbanization and industrialization, had resulted in increasing traffic injuries.

Myanmar was currently engaged in political, economic and social transformation after long period of isolation. Transportation and increased motorization came along with the development of other sectors. At the same time, the estimated number of road traffic deaths escalated 15 per 100000 population,[2] to 20.3 in five years (2011-2015),[1]. It was the second highest death rate in Southeast Asia Region (SEAR) and the utmost peak ever in Myanmar, despite the national target set for 50% reduction of road fatality since 2011,[1].

Within the country, the most common area of RTI was the Nay Pyi Taw,[3], intended area of Myanmar transportation hub in the ASEAN communication network, for both political and economic reason. Since it is a new capital of Myanmar, the government administration officially established there in late 2009. According to 2014 Population and Housing census in Myanmar, Nay Pyi Taw Union Territory was second least populated area, where 2.3% of the country population (1,160,242 people) lived in,[4]. The registered vehicles in this area were only 2.1% of national figures,[5]. However, the highest prevalence and death rate of RTI was seen in this area,[3]. In addition, more than half of the injury cases in public hospitals in Nay Pyi Taw were RTI victims, where one-fifth of the total admitted patients annually,[6].

Beyond the morbidity and mortality statistics, many non-fatal victims were left with short or long-term disabilities with social, psychological and financial burdens,[7]. Accurate and reliable information on the disease burden was required as a baseline data for the policy and decision-makers, before setting the priorities in RTI prevention and control. However, outcome studies focused on the burden of disease approach were very limited for both national and local setting in Myanmar. This study addressed the alarming issue of capital city in Myanmar by quantifying the burden of road traffic injuries in Nay Pyi Taw Union Territory.

## MATERIAL AND METHODS

Cross-sectional study was performed to measure the burden of RTIs in the studied area. All the victims of traffic accidents, both fatal and non-fatal, from injury surveillance registry were analyzed over five-year periods (2012-2016). The patients with incomplete information were excluded from the data set. The burden calculator version 1.10,[8] was used as the statistical tool, which was an open analytical tool for estimating the population burden of injuries. It was Excel-based, including the default set of estimation parameters which were conceptually based on global burden of disease (GBD)-2010 assumption.

Two data sets were added to run the burden calculator. The former included the population counts of the Nay Pyi Taw Union Territory, arranged by age and sex distribution. It was collected from the 2014 census in Myanmar. The latter covered the morbidity and mortality data of traffic injury, which were derived from the injury registry and organized by age, sex, external causes and the outcome of the patients. The age of RTI patients were categorized into 5-years age groups according to GBD guideline. The external causes of injury in international statistical classification of diseases and related health problems, tenth revision (ICD-10) codes were reassigned into health state sequelae by the GBD shortlist of injury code.

The DALYs analysis was performed by three steps; calculating years of life lost due to premature mortality (YLLs), calculating years lived with disability (YLDs) and subsequently summing up those figures into disability-adjusted life years (DALYs). Firstly, YLL was calculated by the following model;

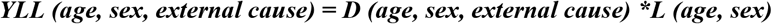

In this formula, D was number of deaths and L was the standard life expectancy at age of death in years. The global standard life table developed for GBD-2010 was used in this study. YLL was mainly calculated by comparing the age of persons who died in transport crashes to the expected life expectancy of that age and gender.

In second step, the incidence of injury conditions, categorized by types of injury or by external causes of injury, were used to estimate the disability or health loss attributable of the non-fatal RTI cases. Since the types of injury were not properly recorded in the primary data set, the external causes of injuries were used, which was applied by the mapping from the external causes to injury sequelae by GBD-2010. The model estimated for YLD was

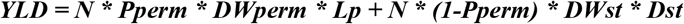

In this formula, N was the number of incident cases of each sequaelae, Pperm was the proportion of cases that had permanent disability, DW was the disability weight that reflected the severity of the health decrement on a scale from 0 (perfect health) to 1 (dead), DWperm was the permanent disability weight, DWst was the short-term disability weight, Dst was the short-term duration of the disabling event, and Lp was the period life expectancy of the population.

In this study, only one injury per victim was counted in the calculations to avoid unrealistically high YLDs for victims. When victims have multiple injuries, the highest YLDs were accepted while the rest were excluded. The disability weight for each injury was applied by GBD-2013.

Finally, the DALY was calculated by summing YLLs and YLDs,

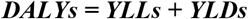

The burden of injuries was analyzed by age, sex and types of road user disaggregation. In the rate calculation, the average numbers was divided by the respective population of Nay Pyi Taw. All of the DALYs, YLLs and YLDs were described by the number in years, as well as the rate per 100,000 population.

## RESULTS

The total of 16,338 RTI patients, including 1015 deaths, was analyzed by sex, age and road user disaggregation. During the study period, the number of RTI cases significantly increased with years, and the amount was double up from 1,820 in 2012 to 3,728 in 2016. Of all the road accidents, male contributed 73.80% of non-fatal and 76.38% of fatal cases. The mean and standard deviation of the age of traffic victims were 31.15 ± 14.57 years. Among the injurers, two out of three were between 15 and 39 years adult. The majority (68.79%) of RTI cases were motorcyclists.

The incidence rate of RTIs in this study was 281.6 per 100,000 population, whereas 427.7 in male and 142.9 in female. The morbidity and mortality rate were 264.1 and 17.5 per 100,000 population respectively, which accounted for the rates of YLD and YLL as 128.1 and 922.8 per 100,000 population. Although the ratio of deaths to alive cases in RTIs was 1:6.6, the rate of YLL was seven times higher than that of YLD.

A total of 60,962 years of life were lost due to traffic crashes, 53,531 (87.81%) of which were related to premature death and the rest, 7,431 (12.19%) were due to disability. On reviewing the average rates of DALYs within studied period (see in table-1), it was 1050.85 per 100,000 population in total with the disaggregation of 808.8 in male and 242.1 in female. Almost entire burden of traffic injuries (92.7%) was contributed by the working age group, 15 to 64 years. Among them, nearly half of burden came from the young adults of 15-29 years. By sex distribution, the highest rate of DALYs in male was seen in the age-group of 20- 24 years while that of female was found in 30-34 years. The detailed descriptions were shown in table (1).

**Table (1).** Average numbers and rates of YLLs, YLDs and DALYs due to RTIs in Nay Pyi Taw Union Territory, by sex and age-group distribution (2012-2016)

| Age-group | YLLs |  |  | YLDs |  |  | DALYs |  |  |
| --- | --- | --- | --- | --- | --- | --- | --- | --- | --- |
|  | Sums* | Percentage | Rate** | Sums* | Percentage | Rate** | Sums* | Percentage | Rate** |
| **Male** |  |  |  |  |  |  |  |  |  |
| <5 year | 136 | 1.64 | 129.73 | 27 | 2.41 | 25.37 | 163 | 1.73 | 155.10 |
| 5-9 year | 126 | 1.52 | 112.87 | 15 | 1.38 | 13.62 | 141 | 1.5 | 126.49 |
| 10-14 year | 162 | 1.96 | 146.98 | 22 | 1.95 | 19.55 | 184 | 1.96 | 166.53 |
| 15-19 year | 1363 | 16.46 | 1342.00 | 132 | 11.96 | 130.18 | 1495 | 15.93 | 1472.18 |
| 20-24 year | 1520 | 18.36 | 1377.18 | 249 | 22.53 | 225.57 | 1769 | 18.85 | 1602.75 |
| 25-29 year | 1403 | 16.94 | 1253.36 | 205 | 18.58 | 183.55 | 1608 | 17.14 | 1436.91 |
| 30-34 year | 1145 | 13.83 | 1135.84 | 143 | 12.90 | 141.51 | 1287 | 13.72 | 1277.35 |
| 35-39 year | 795 | 9.61 | 943.26 | 120 | 10.89 | 142.82 | 916 | 9.76 | 1086.08 |
| 40-44 year | 699 | 8.44 | 943.09 | 64 | 5.77 | 86.10 | 763 | 8.13 | 1029.19 |
| 45-49 year | 347 | 4.19 | 521.56 | 51 | 4.58 | 76.05 | 398 | 4.24 | 597.62 |
| 50-54 year | 257 | 3.10 | 468.51 | 36 | 3.28 | 65.99 | 293 | 3.12 | 534.50 |
| 55-59 year | 205 | 2.47 | 453.27 | 22 | 1.95 | 47.85 | 226 | 2.41 | 501.12 |
| 60-64 year | 61 | 0.74 | 202.91 | 12 | 1.09 | 39.66 | 73 | 0.78 | 242.56 |
| 65-69 year | 42 | 0.52 | 211.40 | 5 | 0.49 | 27.05 | 48 | 0.51 | 238.45 |
| 70-74 year | 3 | 0.04 | 27.51 | 2 | 0.17 | 14.57 | 5 | 0.06 | 42.08 |
| 75-79 year | 8 | 0.09 | 75.61 | 1 | 0.05 | 5.83 | 8 | 0.09 | 81.43 |
| 80-84 year | 4 | 0.05 | 59.60 | 0 | 0.02 | 3.14 | 4 | 0.04 | 62.74 |
| 85+ year | 3 | 0.04 | 67.04 | 0 | 0 | 0.00 | 3 | 0.03 | 67.04 |
| Total | 8279 | 100 | 713.54 | 1106 | 100 | 95.27 | 9384 | 100 | 808.80 |
| **Female** |  |  |  |  |  |  |  |  |  |
| <5 year | 85 | 3.5 | 81.09 | 16 | 4.29 | 15.57 | 101 | 3.61 | 96.66 |
| 5-9 year | 79 | 3.24 | 70.56 | 8 | 2.16 | 7.37 | 87 | 3.10 | 77.92 |
| 10-14 year | 74 | 3.04 | 66.78 | 18 | 4.61 | 15.87 | 91 | 3.25 | 82.65 |
| 15-19 year | 330 | 13.61 | 325.32 | 51 | 13.36 | 50.11 | 382 | 13.58 | 375.43 |
| 20-24 year | 345 | 14.21 | 312.47 | 70 | 18.45 | 63.64 | 415 | 14.78 | 376.11 |
| 25-29 year | 318 | 13.11 | 284.39 | 59 | 15.58 | 53.00 | 378 | 13.44 | 337.39 |
| 30-34 year | 346 | 14.23 | 342.92 | 59 | 15.41 | 58.22 | 404 | 14.39 | 401.14 |
| 35-39 year | 245 | 10.11 | 291.15 | 36 | 9.58 | 43.29 | 282 | 10.04 | 334.44 |
| 40-44 year | 124 | 5.10 | 167.12 | 23 | 6.15 | 31.60 | 147 | 5.24 | 198.72 |
| 45-49 year | 221 | 9.09 | 331.85 | 17 | 4.58 | 26.21 | 238 | 8.48 | 358.06 |
| 50-54 year | 111 | 4.58 | 202.54 | 9 | 2.48 | 17.20 | 121 | 4.29 | 219.74 |
| 55-59 year | 72 | 2.98 | 160.01 | 7 | 1.86 | 15.69 | 79 | 2.82 | 175.70 |
| 60-64 year | 26 | 1.05 | 84.43 | 3 | 0.81 | 10.24 | 29 | 1.02 | 94.67 |
| 65-69 year | 38 | 1.57 | 190.40 | 1 | 0.39 | 7.40 | 39 | 1.41 | 197.79 |
| 70-74 year | 7 | 0.28 | 54.59 | 1 | 0.15 | 4.62 | 7 | 0.26 | 59.21 |
| 75-79 year | 3 | 0.11 | 24.99 | 0 | 0.1 | 3.85 | 3 | 0.11 | 28.84 |
| 80-84 year | 4 | 0.15 | 58.44 | 0 | 0.04 | 1.79 | 4 | 0.14 | 60.22 |
| 85+ year | 1 | 0.04 | 22.57 | 0 | 0 | 0.29 | 1 | 0.04 | 22.86 |
| Total | 2429 | 100 | 209.24 | 378 | 100 | 32.82 | 2808 | 100 | 242.05 |

(\*years) (\*\*per 100,000 population)

The burden of RTI was also investigated by types of road users as shown in table (2). The maximum burden of RTIs was contributed by motorized two-wheelers for all of YLLs, YLDs and DALYs. Although the pattern was similar in both sex, the rate in male was about five times higher than female. The second largest years lost due to early death was seen in pedestrians (9.94% of male and 19.47% of female) and due to disability was seen in car occupants (7.65% of male and 10.59% of female).

**Table (2).** Average numbers and rates of YLLs, YLDs and DALYs due to RTIs in Nay Pyi Taw Union Territory, by sex and road-user distribution (2012-2016)

| Age-group | YLLs |  |  | YLDs |  |  | DALYs |  |  |
| --- | --- | --- | --- | --- | --- | --- | --- | --- | --- |
|  | Sums* | Percentage | Rate** | Sums* | Percentage | Rate** | Sums* | Percentage | Rate** |
| **Male** |  |  |  |  |  |  |  |  |  |
| Pedestrian | 823 | 9.94 | 2250.90 | 59 | 5.34 | 160.13 | 882 | 9.40 | 12055.19 |
| Bicyclist | 140 | 1.69 | 502.75 | 30 | 2.69 | 83.39 | 170 | 1.81 | 2930.70 |
| Motorized 2 Wheeler | 6147 | 74.25 | 13814.66 | 823 | 74.48 | 1868.58 | 6970 | 74.28 | 78416.23 |
| Motorized 3 Wheeler | 38 | 0.46 | 75.96 | 11 | 1.02 | 27.43 | 50 | 0.53 | 516.94 |
| Car | 457 | 5.53 | 1343.56 | 85 | 7.65 | 213.85 | 542 | 5.78 | 7787.03 |
| Bus | 63 | 0.77 | 217.97 | 11 | 0.98 | 26.74 | 74 | 0.79 | 1223.58 |
| Truck | 308 | 3.72 | 734.96 | 40 | 3.64 | 92.95 | 349 | 3.70 | 4139.53 |
| Other | 303 | 3.64 | 760.97 | 46 | 4.20 | 116.60 | 347 | 3.71 | 4387.81 |
| Total | 8279 | 100 | 713.54 | 1106 | 100 | 95.27 | 9384 | 100 | 4043.99 |
| **Female** |  |  |  |  |  |  |  |  |  |
| Pedestrian | 473 | 19.47 | 1153.75 | 34 | 8.84 | 79.52 | 506 | 18.02 | 1233.27 |
| Bicyclist | 78 | 3.20 | 158.35 | 20 | 5.24 | 41.41 | 98 | 3.48 | 199.76 |
| Motorized<br>2 Wheeler | 1259 | 51.83 | 2975.46 | 229 | 60.33 | 486.35 | 1488 | 52.98 | 3461.81 |
| Motorized<br>3 Wheeler | 62 | 2.57 | 157.68 | 7 | 1.89 | 14.93 | 70 | 2.48 | 172.61 |
| Car | 162 | 6.65 | 417.53 | 40 | 10.59 | 93.77 | 202 | 7.20 | 511.30 |
| Bus | 90 | 3.72 | 311.52 | 6 | 1.69 | 13.96 | 96 | 3.44 | 325.48 |
| Truck | 162 | 6.69 | 438.69 | 31 | 8.12 | 69.50 | 193 | 6.88 | 508.19 |
| Other | 143 | 5.87 | 272.69 | 11 | 3.30 | 28.44 | 155 | 5.52 | 301.13 |
| Total | 2429 | 100 | 209.24 | 378 | 100 | 32.82 | 2808 | 100 | 242.05 |

(\*years) (\*\*per 100,000 population)

## DISCUSSION

This is the first study of introducing the burden of disease method into the injury research in Myanmar. The previous burden of diseases studies,[9-11] were related to the Tuberculosis, Dengue and some chronic diseases. Moreover, those studies used the qualitative and descriptive analysis rather than the DAlYs method. In the injury research, the former studies,[12-17] described about the clinical profiles, incidence and its associated factors.

The mortality rate in this study was lower than the WHO estimated traffic fatality rates, but it was two to three times higher than the death rate presented in Myanmar health statistics and police force publications, i.e., 6,[3] and 8.4,[18] per 100,000 population respectively. In addition, the incidence of traffic injury in this study was 281.6 per 100,000 population, which was also higher than that from Myanmar health statistics; i.e., 155 per 100,000 population,[18]. It was due to the different data definition and natures of data collection purposes. By comparing with other countries in Asia Highway Network, the similar mortality rate was seen as 17.7 in Cambodia, 16.6 in India and 13.6 in Bangladesh,[1].

Although fatal cases were only 6.2% in the study, it attributed 87.8% of the total burden. The same finding was found in the global burden of injury study,[5], and the studies from developing countries; accounting of YLL in total burden as 75% in Nepal, 88% in Thailand, 87% in Iran and 90% in Brazil respectively,[19-22]. All of these findings indicated that death numbers highly influenced in the burden of traffic injury even in different countries. Moreover, the major contribution of RTI burden in this study came from the premature death, not from the disability. It was agreed with the GBD analysis for Myanmar in 2016, in which the burden of traffic injuries was seen in all ages by YLLs, however, it was not a prioritized problem for the disability by analyzing its YLD,[23].

This study also highlighted the sex and age differences. The half of the burden in this study came from young and active age group of 15 to 29 years. In the other studies, the highest YLD was found in the age group 15-29 in the Iran,[24] and 15-24 in the France,[25]. It indicated the vulnerability of youth in traffic accidents. In the DALY approach, the younger the age of RTI patient, the greater burden in this community. The similar finding was seen in the GBD profile for Myanmar,[23], in which described that road injuries took eighth leading causes of death for all ages, however, by calculating the years of life lost, it jumped up to the third position.

In this study, male contribution was higher in all age groups and generally 77% of total RTI burden. The mortality of RTI in male was three to five times higher than female. This finding was in line with the latest global status report on road safety which described that men attributed three out of four road deaths globally,[6]. In Myanmar culture, males were designated to be head of the household and led the socioeconomic of their families: therefore, their health burden was directly affected to their families. The dominance of male involvement in RTI burden was consistently seen with other RTI studies,[20, 24-25] albeit the figure varied,

The greatest burden of RTI came from the motorcyclists in both sex and all age-groups. Such kind of disproportionate burden was influenced by many factors. Public transportation within the city was very limited. Due to its convenient and affordable price, motorcycle became the main choice of transportation in for both urban and rural community. More than three-quarter (76.2%) of registered vehicles in Nay Pyi Taw,[5] were motorcycles, However, the risky behaviors of the RTI victims, e.g., high speed, lack of wearing helmets, were not included in this study.

## CONCLUSIONS

This study also demonstrated the potential of using local data from injury registry for apply the burden of disease concept. The findings quantified the burden in the community and addressed the major risk factors of traffic injuries in such areas. It was unprecedentedly initiated as the baseline data for the future investigations as well as the very first input of DALY method advocated for the local policy makers. Similar methodological studies in the same or different areas were suggested for addressing comparability and facilitating improved data quality. The unique data banks for injury surveillance, in turn, required strong commitment and collaborations of concerned parties at all levels.

## Data Availability

The data that support the findings of this study are available on request from the corresponding author, email.

## ETHICAL APPROVAL

This study was approved by the Institutional Review Board (IRB) of the Faculty of Public Health, Mahidol University, IRB number – 116/2559, COA. No. MUPH 2016-146, and Ethics Review Committee of the Ministry of Health and Sports, Myanmar, ERC number-012416/Ethics/DMR/2016/144.

## ACKNOWLEDGEMENT

This work was completed as part of the Dr.PH program in global health, Faculty of Public Health, Mahidol University, Thailand.

## COMPLETING INTERESTS

No completing interests

